# Investigating the marginal and herd effects of COVID-19 vaccination for reducing case fatality rate: Evidence from the United States

**DOI:** 10.1101/2023.03.11.23287133

**Authors:** Tenglong Li, Zilong Wang, Shuyue He, Ying Chen

## Abstract

Vaccination campaigns have been rolled out in most countries to increase the vaccination coverage and protect against case mortality during the ongoing pandemic. To evaluate the effectiveness of COVID-19 vaccination, it is vital to disentangle the herd effect from the marginal effect and parameterize them separately in a model. To demonstrate this, we study the relationship between the COVID-19 vaccination coverage and case fatality rate (CFR) based on a U.S. vaccination coverage at county level, with daily records from March 11^th^, 2021 to Jan 26^th^, 2022 for 3109 U.S. counties. Using segmented regression, we discovered three breakpoints of the vaccination coverage, at which the herd effects could potentially exist. Controlling for county heterogeneity, we found the size of the marginal effect was not constant but actually enlarged as the vaccination coverage increased, and only the herd effect at the first breakpoint was statistically significant, which implied indirect benefit of vaccination may exist at the early stage of a vaccination campaign. Our results have demonstrated that public health researchers should carefully differentiate and quantify the herd and marginal effects in analyzing vaccination data, to better inform vaccination campaign strategies as well as evaluate vaccination effectiveness.

## 1 Introduction

The world has been living in the tunnel of COVID-19 pandemic since the outbreak in 2019, yet without a clear idea about its outlet. A great hope has been placed in COVID-19 vaccines to end the pandemic, as clinical trial results suggested COVID-19 vaccination can effectively prevent symptomatic infections especially severe symptoms, which protects against mortality associated with infections [1-3]. For this reason, public demand for COVID-19 vaccines was fervent and vaccination campaigns were initiated all over the world, for an early safe vaccine supply for populations at risk as well as a massive vaccine supply to match the public’s demand [4-5]. For an example, the Food and Drug Administration (FDA) issued emergency use authorizations (EUA) for Pfizer-BioNTech and Moderna COVID-19 vaccines in December 2020, which marked the beginning of the vaccination campaign in the U.S. COVID-19 vaccines were then first allocated for populations at risk, the elderly population (age 65+) and the frontline (mostly healthcare and education) workers. After president Biden announced that all Americans would be eligible for COVID-19 vaccines by May 1^st^, 2021, the vaccination campaign was further accelerated [6]. Booster doses of COVID-19 vaccines were introduced to restore the level of protection (antibody) eroded by time [7-9]. By November 24, 2022, more than 80% of Americans have received at least one dose and more than 68% of Americans have completed a primary series of COVID-19 vaccine [10]. Literature has reported that the vaccination coverage is negatively associated with case fatality rate (CFR), which refers to the mortality rate among those who are infected (i.e., confirmed COVID cases) [11-12].

It is necessary to decompose the protection effect of COVID-19 vaccines in order to better understand the underlying mechanism [13]. The protection effect of COVID-19 vaccines is in general a mix of two different effects, i.e., the direct effect and the indirect effect [14]. The direct effect refers to direct protection of inoculated individuals, as the vaccines can effectively reduce individual susceptibility to COVID-19 infection and severe symptoms [13-14]. The indirect effect, however, is a bit abstract and attributed to herd immunity, which is a conception states that transmission of the agent can be largely prevented if a fixed proportion of the population is immunized (either by vaccination or by recovery from infection; this proportion is called herd immunity threshold), rendering an infectious disease insignificantly dangerous for public health [15-16]. The indirect effect is defined as the protection gained by unvaccinated people, through the reduced number of infected people in the population as well as their reduced infectiousness, which can be achieved by vaccinating certain proportions of the population [13]. It should be noted here that those proportions we mentioned above are different from herd immunity threshold as they potentially correspond to different levels of herd immunity in a population [17]. In fact, those proportions are thresholds for triggering the indirect effect (with different sizes) in the course of a vaccination campaign for a target population.

The above concepts of the direct and indirect effects should be contextualized in the investigation of the impact of COVID-19 vaccination on case fatality rate (CFR). The direct effect could be interpreted as the reduction in CFR associated with one unit/percent increase in the vaccination coverage, i.e., the direct effect evaluates the marginal gain during a vaccination campaign. For this reason, the direct effect is referred to as the marginal effect in this paper. The indirect effect could be interpreted as the additional reduction in CFR if the vaccination coverage passes certain unknown thresholds, i.e., the indirect effect quantifies the additional gain potentially due to herd immunity in the process of vaccinating a target population. To better characterize its nature, the indirect effect is referred to as the herd effect in this paper. It’s particularly important to disentangle the herd effect from the marginal effect, for the following three reasons: First, the marginal and herd effects address different scientific questions with regard to distinct groups of people (i.e., the vaccinated individuals versus the unvaccinated individuals). Second, as discussed earlier, there are underlying thresholds for triggering the herd effects, and those thresholds essentially delineate different stages in a vaccination campaign where the marginal and herd effects may not be constant across those stages. Third, given the aforementioned two reasons, a deeper knowledge about the protection effect of vaccination is likely gained by learning the marginal and herd effects, and vaccination strategies could be optimized for a target population based on such knowledge. Unfortunately, we haven’t seen research on this important topic so far. Our goal in this paper is to estimate the herd and marginal effects based on a dataset from the U.S. Centers for Disease Control and Prevention (CDC), which records various vaccination coverages for each U.S. county daily [10]. We hypothesize that both the herd and marginal effects exist and are significantly negative for modeling CFR. The segmented regression is employed first to identify the breakpoints which are considered as the thresholds for triggering the herd effect, based on data of all the U.S. counties included in our study. With the identified breakpoints, we estimate the herd and marginal effects at national level using segmented regression and at county level using mixed model. Data on social vulnerability index (SVI) for individual counties is also included to control for health disparities due to sociodemographic factors at county level [18]. Heterogeneity among individual counties is further evaluated by the random effects associated with the herd and marginal effects among in a mixed model.

This paper is structured as follows: In the next section of materials and methods, the data used in this paper will be described in details, along with the models adopted for analyses at both national level and county level. The results from our analyses at national level and county level are presented and explained in the section of results, with a focus on the estimation and interpretation of the herd and marginal effects of vaccination regarding CFR. Our findings will be summarized in the discussion section, where important implications and limitations of our study will also be discussed.

## 2 Materials and Methods

### 2.1 Data

Our data comes from three different sources. The US vaccine administration and equity dataset is obtained from the CDC website and has vaccination coverages of the general population and its subpopulations (defined by age) recorded daily at county level [10]. The percent of people who completed a primary series of vaccination in the general population was extracted from the dataset and served as the main covariate in our model. The daily CFR at county level was calculated as the ratio between the daily count of deaths and the daily count of COVID-19 cases, based on the time series summary tables of COVID-19 deaths and confirmed cases, which were accessed from the COVID-19 data repository by the Center for Systems Science and Engineering (CSSE) at John Hopkins University [19]. To further control for county heterogeneity, we used a dataset from the Centers for Disease Control and Prevention Social Vulnerability Index (CDC SVI) database, created by the Geospatial research, Analysis & Services Program under the Agency for Toxic Substances and Disease Registry [18]. The CDC SVI database was established to help health officials and emergency response planners identify counties that will most likely need support before, during, and after a hazardous event. CDC SVI ranks counties on 15 social factors and further groups them into four themes, namely socioeconomic status, household composition & disability, minority status & language, and housing type & transportation [20]. We chose to use the theme-specific ranking which was constructed by summing the percentiles of the factors under each theme. The theme-specific ranking was set in the range from 0 to 1, with higher values indicating greater vulnerability.

The vaccination coverages and daily CFR for the period between March 11^th^, 2021 and Jan 26^th^, 2022 were selected. We chose March 11^th^, 2021 as it was the date when president Biden announced that COVID-19 vaccine would be available for all American adults by May 1^st^, 2021, an event marked the beginning of massive vaccination campaign in the U.S. We chose Jan 26^th^, 2022 as the ending date of our study as it was reported on this date that Omicron variant accounted for 99.9% of the new infections. This would alleviate the concern of potential confounding effect of Omicron variant regarding the relationship between the vaccination coverage and CFR. 31 counties with missing values on county FIPS code, vaccination coverages, the CDC SVI or CFR were excluded, and the final dataset has 1001098 observations clustered by 3109 U.S. counties. To prepare the dataset for analysis at national level, we further extracted the average CFR and average vaccination coverage (i.e., the percent of people who completed a primary series of COVID-19 vaccine) across all the counties in our dataset for each day during our study period.

### 2.2 Models

Segmented regression models were employed to estimate the herd and marginal effects. Segmented regression is very similar to ordinary regression, with the only difference that regression coefficients should be estimated repeatedly for different local regions whose boundaries are defined by breakpoints, which represent the thresholds of structural changes in regression models [21-22]. Typically, the first step is to determine the number of breakpoints, which can be achieved by a model selection alike procedure, i.e., models with different number of breakpoints are compared in terms of their model fit indices (such as AIC or BIC) to determine the optimized number of breakpoints. The second step is to estimate the locations of breakpoints given the number of breakpoints. The third step, based on the estimated breakpoints, is then to fit regression models to different local regions separated by the breakpoints. Normally, one would expect all regression coefficients to be changeable across different regions, unless otherwise specified. For our analysis at national level, we intend to examine the relationship between CFR and the vaccination coverage, based on the dataset comprising only the average vaccination coverage and CFR in the U.S. The following regression model is formulated for the analysis at national level:

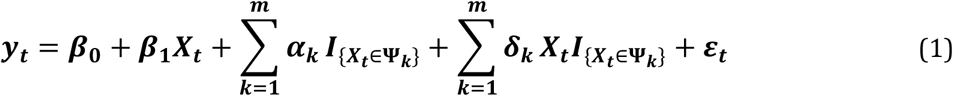

where *y*_*t*_and *X*_*t*_ denote the average CFR and vaccinate coverage in the U.S on day *t*. The model (1) is built on the estimated breakpoints *b*_1_ < *b*_2_ < ⋯ < *b*_*m*_, which implies there are *m* + 1 different local regions and *m* different breakpoints in total (except *b*_0_ and *b*_*m*+1_which are the minimum and maximum of *X*_*t*_). The local regions separated by the breakpoints are denoted by Ψ_*k*_ = [*b*_*k*_, *b*_*k*+1_) for *k* = 1, 2, ⋯, *m*. The reference local region Ψ_0_, although omitted from the model (1), refers to the local region Ψ_0_ = [*b*_0_, *b*_1_). The indicator function 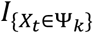 creates the dummy variable which assigns value 1 if the value of *X*_*t*_ falls in the local region Ψ_*k*_ and 0 otherwise, which operationally divides the range of *X*_*t*_ into the local regions. The marginal effects in those local regions are characterized by *β*_1_ for the reference region and *β*_1_ + δ_*k*_ for the local region Ψ_*k*_, and these parameters quantify the marginal gain/drop in CFR if the vaccination coverage increases by one percent. The herd effects for the local region Ψ_*k*_ relative to its previous region are characterized by *α*_*k*_− *α*_*k*−1_(for Ψ_1_ it is just *α*_1_), as *α*_*k*_ quantifies the additional gain/drop in CFR if the vaccination coverage passes the threshold *b*_*k*_, compared to the intercept term *β*_0_ in the reference region Ψ_0_.

The breakpoints *b*_*k*_ *k* = 1, 2, ⋯, *m*, are estimated based on the model (1) and the dataset for the analysis at national level (i.e., with only average daily CFR and vaccination rate in the U.S.). Naturally, they reflect the structural changes in the relationship between CFR and the vaccination coverage in general, and they can be applied to the analysis at county level where we use the longitudinal data (322 days) for all the counties (3109 counties), along with the CDC SVI indicators for explaining county heterogeneity. We build the following mixed model for the analysis at county level:

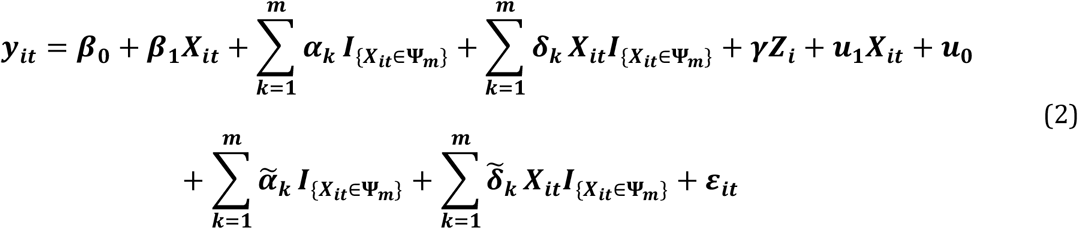

where *y*_*it*_ and *X*_*it*_ denote the CFR and vaccination coverage for county *i* at day *t. Z*_*i*_ is the covariate vector that contains CDC SVI theme-specific rankings on the four main themes for county *i*. *β*_0_, *β*_1_, *α*_*k*_, δ_*k*_ are the parameters characterize the herd and marginal effects, as similarly defined in the model (1), except in the model (2) they are fixed effects. Correspondingly, we have their random effects characterized by *u*_0_, *u*_1_, 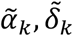, that are due to the heterogeneity among the counties that cannot be explained away by the fixed effects of county rankings on CDC SVI, which are represented by γ. The Model (2) is built on the same set of breakpoints *b*_*k*_, *k* = 1, 2, ⋯, *m*, that are obtained based on the model (1) and the dataset for the analysis at national level. This means the model (2) shares the same local regions Ψ_*k*_ = [*b*_*k*_, *b*_*k*+1_) for *k* = 1, 2, ⋯, *m*, across all the counties in our study. The significances of the fixed effects *β*_0*i*_, *β*_1*i*_, *α*_*k*_, δ_*k*_ as well as their corresponding random effects *u*_0_, *u*_1_, 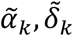, will be checked via model outputs and comparison tests.

## 3 Results

### 3.1 The results of the analysis at national level

As mentioned above, the dataset used for the analysis at national level has two variables, i.e., average daily CFR and average vaccination coverage in the U.S.. The breakpoints were estimated based on this dataset using the “segmented” package in R (version 4.2.0) [22]. To avoid overfitting, we set the maximum number of the breakpoints as 3, based on the curve between the average daily CFR and the average daily vaccination coverage depicted in Figure 1. The segmented package then does an automatic selection of the number of breakpoints based on BIC, and it estimated the locations of the breakpoints conditional on the optimized number of the breakpoints. The estimated breakpoints were superimposed on the curve in Figure 1, to further validate those estimates align with the observed structural changes.

**Figure 1.**
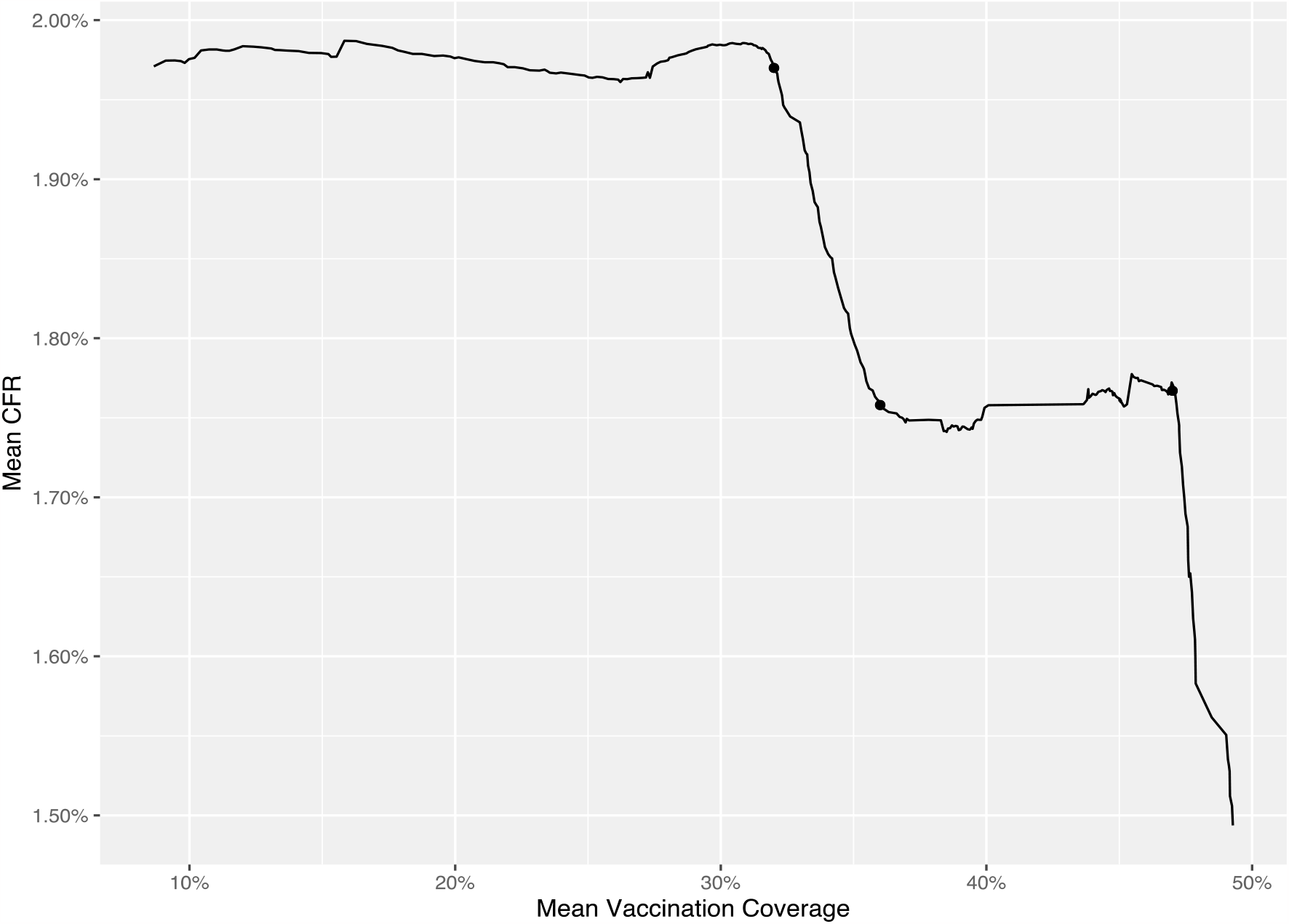
The relationship between the average vaccination coverage and the average CFR in the U.S. The solid dots represent the breakpoints estimated by the “segmented” package in R.

The breakpoints were estimated as 32%, 36% and 47%, which suggested that the herd effect of vaccination may be associated with the thresholds of 32%, 36% and 47% in the vaccination coverage. Based on those breakpoints, we have four different local regions, namely Ψ_1_ = [8.66%, 32%); Ψ_2_ = [32%, 36%); Ψ_3_ = [36%, 47%); Ψ_4_= [47%, 49.28%), with the minimum and maximum of the average daily vaccination coverages as 8.66% and 49.28% respectively. Table 1 lists the estimates of the regression coefficients based on the model (1). We further calculated the herd and marginal effect estimates which are tabulated in Table 2. The marginal effect in the first local region Ψ_1_ was insignificant, which suggested that the drop in the CFR per percent increase in the vaccination coverage was not significantly different from 0, if the vaccination coverage did not surpass 32%. The herd effect at the threshold 32% was also insignificant, which was largely due to the insignificant marginal effect in the region Ψ_1_. We found a significant marginal effect in the second local region Ψ_2_ (−0.057), which indicated that there was a drop of 0.057 percent in the CFR for every percent increase in the vaccination coverage in this region, evidencing that the protection effect of COVID vaccination against mortality. In addition, the herd effect at the threshold 36% was significant too (− 0.233), suggesting that there was a further drop of 0.233 percent in the CFR besides the marginal CFR reduction per percent increase in the vaccination coverage. In the third local region Ψ_3_, however, we observed a slight positive marginal effect in the CFR (0.003), which means the marginal gain of vaccination (in terms of the reduction in CFR) disappeared and vaccination was somehow harmful for protecting against mortality. Correspondingly, the herd effect at the threshold 47% was also positively significant (0.009), suggesting again that vaccination was not helpful at this stage. The marginal effect in the fourth local region Ψ_4_ was strongly negative, specifically there was a drop of 0.115 percent in the CFR associated with every percent increase in the vaccination coverage at this stage.

**Table 1.** The regression model parameter estimates for the analysis at national level.

| Parameter | Estimate | T Ratio | p-value |
| --- | --- | --- | --- |
| $\beta_0$ | 1.976 | 624.70 | <0.001 |
| $\beta_1$ | 0.000 | 0.28 | 0.78 |
| $\alpha_1$ | -0.003 | -0.90 | 0.37 |
| $\delta_1$ | -0.057 | -45.32 | <0.001 |
| $\alpha_2$ | -0.236 | -80.67 | <0.001 |
| $\delta_2$ | 0.003 | 7.50 | <0.001 |
| $\alpha_3$ | -0.227 | -55.09 | <0.001 |
| $\delta_3$ | -0.115 | -47.03 | <0.001 |

**Table 2.**
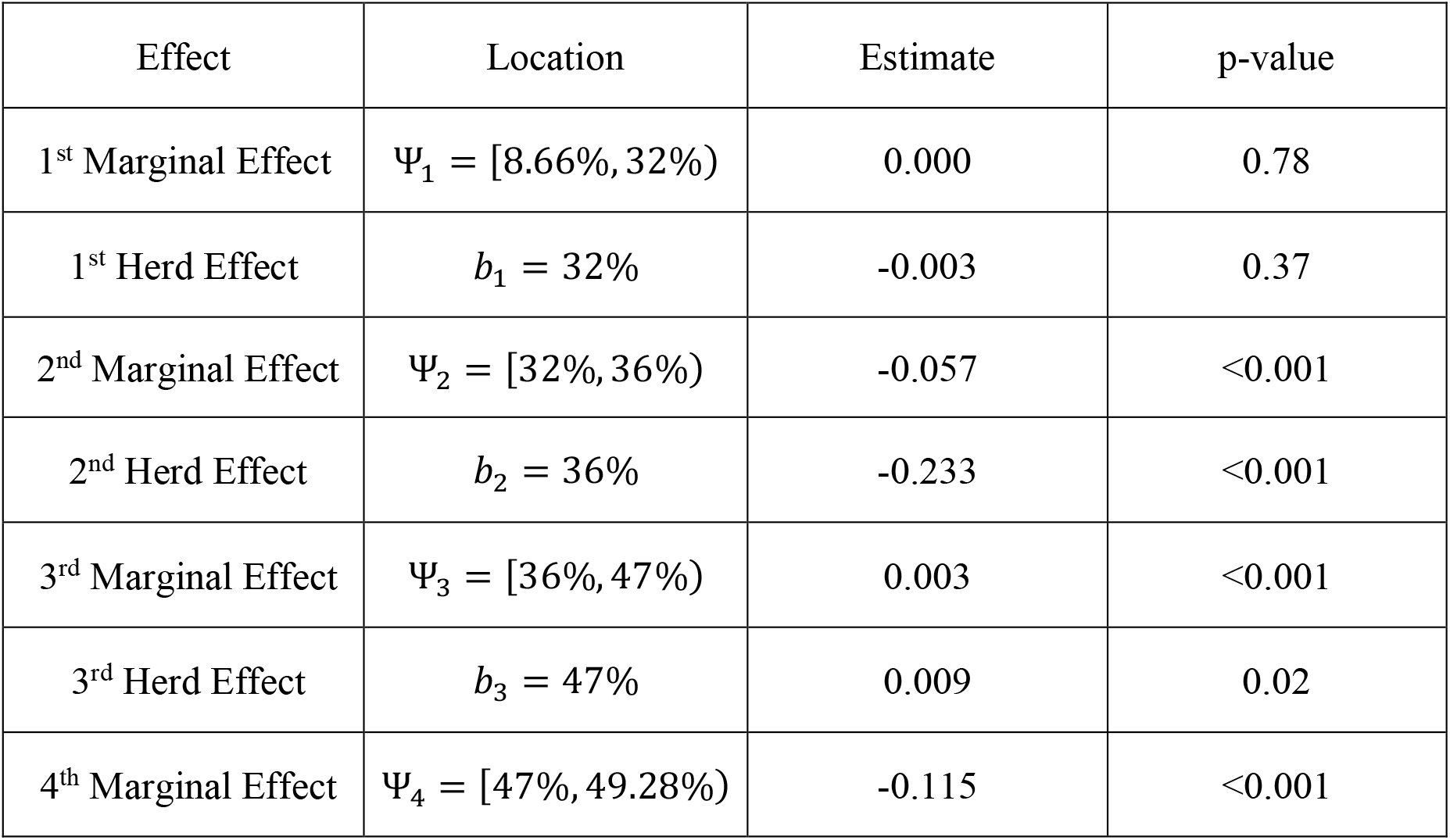
The herd and marginal effect estimates for the analysis at national level.

### 3.2 The results of the analysis at county level

We further investigated the marginal and herd effects of COVID vaccination based on an analysis at county level, where the daily CFR and vaccination coverages from March 11^th^, 2021 to Jan 26^th^, 2022 as well as the CDC SVI rankings for 3109 U.S. counties were used. The estimated breakpoints of 32%, 36% and 47%, obtained based on the analysis at national level, were adopted for our analysis at county level. The mixed model (2) was employed to account for the clustered data at county level, and its fixed and random effect estimates are tabulated in the Table 3. Furthermore, the estimates of herd and marginal effects, as well as their corresponding random effect estimates, are listed in the Table 4. To determine the significance of the random effects, we compared the full model (i.e., the model (2)) with two different reduced models (one without the random effects associated with all the marginal effects, i.e., *u*_1_, 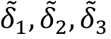; another one without the random effect associated with the first marginal effect only, i.e., *u*_1_), and the resultant tests gave p-values smaller than 0.001, suggesting that it was necessary to include random effects for all the marginal and herd effect parameters.

**Table 3.** The mixed model parameter estimates pertaining to fixed effects (F.E.) and random effects (R.E.) for the analysis at county level. The standard errors (S.E.) of the random effects (R.E.) are also provided.

| Parameter | F.E. Est | F.E. T Ratio | F.E. p-value | R.E. Est | S.E. of R.E. |
| --- | --- | --- | --- | --- | --- |
| $\beta_0$ | 1.64 | 28.88 | <0.001 | 1.33 | 0.03 |
| $\beta_1$ | -0.004 | -8.85 | <0.001 | 0.0005 | 0.00001 |
| $\alpha_1$ | -0.025 | -7.35 | <0.001 | 0.024 | 0.0008 |
| $\delta_1$ | -0.007 | -3.74 | <0.001 | 0.007 | 0.0002 |
| $\alpha_2$ | -0.018 | -2.84 | 0.005 | 0.091 | 0.003 |
| $\delta_2$ | -0.02 | -13.07 | <0.001 | 0.006 | 0.0002 |
| $\alpha_3$ | -0.004 | -0.35 | 0.728 | 0.2 | 0.007 |
| $\delta_3$ | -0.039 | -23.49 | <0.001 | 0.004 | 0.0002 |
| $\gamma_1$ | 0.0004 | 0.33 | 0.744 | n/a | n/a |
| $\gamma_2$ | 0.807 | 11.00 | <0.001 | n/a | n/a |
| $\gamma_3$ | -0.057 | -0.73 | 0.467 | n/a | n/a |
| $\gamma_4$ | 0.004 | 0.05 | 0.961 | n/a | n/a |

**Table 4.**
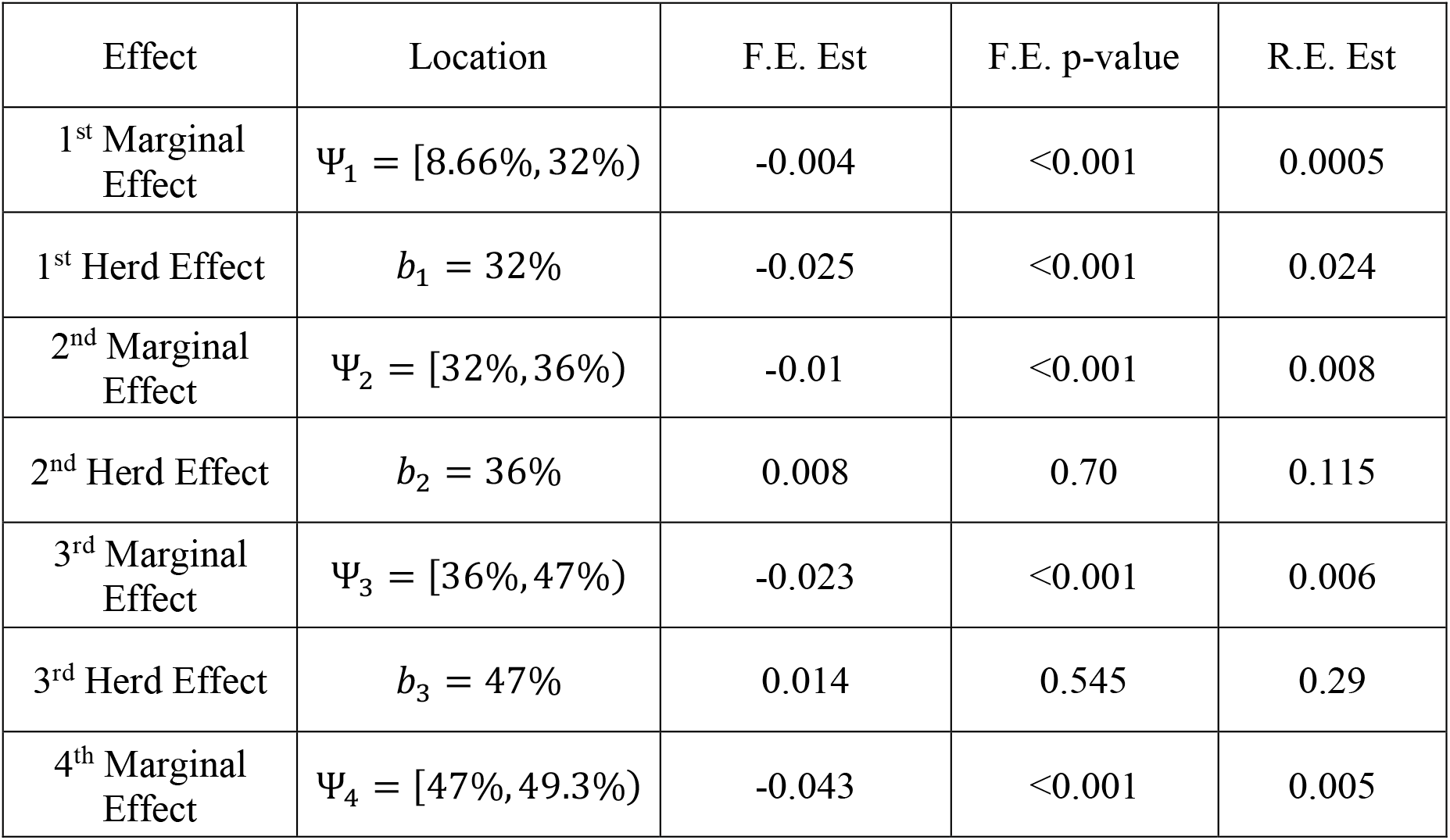
The herd and marginal effect estimates for the analysis at county level. The fixed effect estimates (F.E. Est), the random effect estimates (R.E. Est) as well as the p-value for the F.E Est are provided.

Across all the U.S. counties in our data, the marginal effect was significantly negative in the first local region Ψ_1_ (i.e., when the vaccination coverage was between 8.66% and 32%), specifically one percent increase in the vaccination coverage was associated with 0.004 percent drop in the CFR. The first herd effect at the threshold of 32% was -0.025 and significant, meaning there was an additional drop of 0.025 percent in the CFR as the vaccination coverage reached 32%, beyond the marginal effect observed in Ψ_1_. The marginal effect in the second local region Ψ_2_ was also significantly negative (−0.01), which showed that there was 0.01 percent drop in the CFR per one percent increase in the vaccination coverage, when the vaccination coverage was between 32% and 36%. The second herd effect, however, was overall insignificant, which suggested additional protection effect at the threshold 36% may not exist. Similarly, we found significant marginal effect (−0.023) for the third local region Ψ_3_ but insignificant herd effect at the threshold 47%. The marginal effect within the fourth local region Ψ_4_ was the strongest, as every percent increase in the vaccination coverage was associated with 0.043 percent reduction in the CFR, if the vaccination coverage surpassed 47%.

Furthermore, heterogeneity among the U.S. counties regarding the herd and marginal effect estimates was evident. For the first herd effect (at 32%), the fixed effect estimate was -0.025 with a random effect of 0.024, and this means roughly 56% of the counties had negative herd effects as expected, but the other 44% of the counties could have no herd effect or even positive herd effects at the threshold 32%. For the second and third herd effect (at 36% and 47% respectively), roughly 49% of the counties have negative herd effects, which further demonstrated that those two herd effects were not significant among the counties. Regarding the marginal effects: although their fixed effect estimates were all very significant (p-value < 0.001), their random effect estimates suggested the fourth marginal effect was the strongest one (was negative in 73% of the counties). The first, second and third marginal effects were negative in approximately 57%, 54% and 62% of the U.S. counties. All taken, the protection effect of COVID vaccination was confirmed in general and for the majority of the U.S. counties, while substantial heterogeneity that defined the size and the validity of the protection effect for individual county still existed. We also found that only one CDC SVI theme ranking, i.e., rankings on household composition & disability, could help explain the county heterogeneity. Unsurprisingly, this CDC SVI theme ranking was positively related to CFR, and specifically one percentile rise in the theme ranking could result in 0.8 percent increase in CFR.

## 4 Discussion

Vaccination has been acknowledged as an effective tool to reduce hospitalization and mortality related to COVID-19 infections, and vaccination campaign has been rolled out in virtually every country that has access to COVID-19 vaccines. Understanding the effect of COVID-19 vaccination in terms of case fatality rate (CFR) reduction has unquestionably profound meaning, for a successful implementation of the COVID-19 vaccination campaign. Drawing on the direct and indirect effects of vaccination from literature, we rename the direct effect as the marginal effect of vaccination and the indirect effect as the herd effect of vaccination, to better describe the nature of those effects in terms of reducing the CFR. Defining the herd and marginal effects also helps build regression models for obtaining their estimates, as those two kinds of effects require different parameterization in the model. Analysis at the national level and county level for the United States, were then implemented based on datasets containing the daily vaccination coverages and case reports in the U.S. Theme rankings for individual counties from the CDC SVI were also included to explain heterogeneity at county level. Our analysis at national level suggested three different locations (i.e., when the vaccination rate reached 32%, 36% and 47%) for possible herd effects and strong significance for the marginal effects, which was further confirmed by our analysis at county level after controlling for county heterogeneity.

Our analyses have demonstrated how COVID-19 vaccination protects against COVID related mortality over the course of COVID-19 vaccination campaign in the U.S. In general, COVID-19 vaccination indeed can significantly reduce the CFR, but its effect is not constant during the vaccination campaign. The estimated breakpoints have divided the vaccination campaign into four different regions based on the vaccination coverage, i.e., Ψ_1_ = [8.66%, 32%), Ψ_2_ = [32%, 36%), Ψ_3_ = [36%, 47%) and Ψ_4_ = [47%, 49.3%). The marginal effects in those four regions are correspondingly -0.004, -0.01, -0.023 and -0.043, which are all significant. This shows the vaccination can directly result in meaningful reduction in the CFR and thus it should be recommended especially for the unvaccinated population, as the marginal effects largely quantify the reduced risks of mortality that one would benefit from the vaccination if he/she chooses to get vaccinated. We also observe that the sizes of the marginal effects enlarge as the vaccination coverages increases, which suggests that the direct benefit of COVID-19 vaccination is becoming more and more significant as the vaccination coverage in the population increases. Our results also indicate the existence of herd effect, specifically at the threshold 32%. The herd effect at the threshold 32% is statistically significant (−0.025), which demonstrates the indirect (additional) benefit brought by the vaccination once the vaccination coverage reaches 32% in the population. This implies that one would indirectly benefit from the COVID-19 vaccination even if he/she is not vaccinated as long as the vaccination coverage passes 32%, by a 0.025% reduction in the CFR.

Our results have important implications for the COVID-19 vaccination strategies. First, our findings suggest that vaccination campaign should be rapidly carried out at the initial stage, to trigger the threshold for herd effects, in order to procure additional protection of COVID-19 vaccination against the CFR for the entire population regardless of individual vaccination statuses. This echoes our earlier finding of the significant herd effect at the first breakpoint 32% and is consistent with recommendations offered by the literature [3,4,23,24,25]. It is noteworthy that, a rapid effective implementation at the initial stage can pose considerable logistical challenges for a vaccination campaign [23,26,27].

Therefore, careful resource planning is required for the access, transportation, storage and distribution of vaccines, which has been exemplified by the vaccination campaign in the U.S. [25,28]. Second, eligible unvaccinated individuals should be encouraged (even urged) to get vaccinated at all stages of a vaccination campaign, as the marginal effects were evident across all the local regions defined for the U.S. vaccination campaign in our analysis. More profoundly, we found the whole population would benefit more if more people got vaccinated, as the size of marginal effect was positively correlated with the vaccination coverage in the population. The gain from the marginal effects, on average, also outweighed the gain from the herd effects, as manifested by the Table 4. These key observations suggest that the marginal effect is more important than the herd effect for the protection against COVID mortality [15]. Thus, vaccination strategy should focus on how to capitalize on the marginal effect, i.e., promote individual vaccination willingness and accessibility, in order to continuously push for a higher vaccination rate in the population [15,29]. Based on our results, the goal of a vaccination campaign should be pursuing a higher vaccination coverage in the population, rather than meeting a predefined threshold for triggering the herd effect [4,29,30].

Heterogeneity among the U.S. counties in terms of the marginal and herd effects is considerable. The sizes and even the signs of the marginal and herd effects could vary across all the counties, which signals that the protection effect of COVID-19 vaccination is not constant and partially determined by county idiosyncrasy. For example, we took the social vulnerability index (SVI) into account in our analysis and did find the theme of household composition & disability significantly was significantly associated with the CFR after controlling for the vaccination coverage. This indicates that demographical features of individual county, such as the age distribution and disability proportion, play vital roles in explaining the heterogeneity existed for the relationship between the vaccination coverage and the CFR [31]. Although the other three SVI themes, namely socioeconomic status, minority status & language and housing type & transportation, were not statistically significant, factors such as environmental conditions [32], political atmosphere [33] and non-pharmaceutical interventions [34] could contribute to county heterogeneity, and potentially confound the relationship between the vaccination coverage and the CFR. Most notably, research has shown that vaccine hesitancy (willingness) is a key determinant of vaccination coverage, and it potentially mediates the relationship between the factors influencing the CFR (like SVI) and the CFR itself, and therefore variation of vaccine hesitancy among the U.S. counties potentially accounts for a significant portion of the county heterogeneity observed in our paper [35-36].

There are limitations in our analysis: We did not investigate the impact of COVID-19 variants on the CFR and the vaccine effectiveness, considering there were different COVID-19 variants (and their lineages and sublineages), such as alpha, delta and omicron, spreading during our study period, as we can hardly identify the boundaries of the spreading period of each variant from the data. For the similar reason, the potential impact of different brands of vaccines (such as BioNTech and Moderna) was also not considered in our model, as the data did not contain information about the number of administered doses of every specific brand. Most importantly, our model treats the breakpoints as the fixed values across all the counties, which may not be true as the breakpoints could vary across different counties as a result of unique evolvement of vaccination campaign in individual counties. Unfortunately, allowing each county to have its own breakpoints would require a huge number of parameters and a complex Bayesian model, which goes beyond the scope of this paper [37]. Therefore, further robustness and sensitivity analyses may be warranted [38-41].

To summarize, we have shown the existence of the herd effects via a segmented regression model. Specifically, we identified three different breakpoints that represented the locations of the herd effects. Accounting for county heterogeneity, we found one of the three herd effects to be statistically significant, and it suggested that additional indirect benefit of COVID-19 vaccination may exist at the earlier stage of a vaccination campaign. We also found the marginal effect size varied at different stages of the vaccination campaign, and specifically the marginal (direct) benefit of COVID-19 vaccination likely became larger as the vaccination coverage increased. Our findings demonstrate that the herd and marginal effects should be carefully differentiated and assessed in analyzing vaccination data, to better inform vaccination campaign strategies as well as evaluate vaccination effectiveness.

## Data Availability

All data produced are available online at the following websites:

https://www.atsdr.cdc.gov/placeandhealth/svi/data_documentation_download.html

https://covid.cdc.gov/covid-data-tracker/#vaccine-delivery-coverage

https://github.com/CSSEGISandData/COVID-19

## References

[1] Dal-Ré R, Bekker LG, Gluud C, et al. Ongoing and future COVID-19 vaccine clinical trials: challenges and opportunities. Lancet Infect Dis. 2021;21(11):e342–e347. doi:10.1016/S1473-3099(21)00263-2

[2] Yan ZP, Yang M, Lai CL. COVID-19 vaccines: A review of the safety and efficacy of current clinical trials. Pharmaceuticals (Basel). 2021;14(5). doi:10.3390/ph14050406

[3] Ahmed S, Khan S, Imran I, et al. Vaccine development against COVID-19: Study from pre-clinical phases to clinical trials and global use. Vaccines (Basel). 2021;9(8):836. doi:10.3390/vaccines9080836

[4] World Health Organization. Global Covid-19 Vaccination Strategy in a Changing World; 2022. https://www.who.int/publications/m/item/global-covid-19-vaccination-strategy-in-a-changing-world--july-2022-update

[5] Mathieu E, Ritchie H, Ortiz-Ospina E, et al. A global database of COVID-19 vaccinations. Nat Hum Behav. 2021;5(7):947–953. doi:10.1038/s41562-021-01122-8

[6] Hodge JG. Nationalizing public health emergency legal responses. J Law Med Ethics. 2021;49(2):315–320. doi:10.1017/jme.2021.45

[7] Bar-On YM, Goldberg Y, Mandel M, et al. Protection of BNT162b2 vaccine booster against Covid-19 in Israel. N Engl J Med. 2021;385(15):1393–1400. doi:10.1056/NEJMoa2114255

[8] Mahase E. Covid-19 booster vaccines: What we know and who’s doing what. BMJ. 2021;374:n2082. doi:10.1136/bmj.n2082

[9] Andrews N, Stowe J, Kirsebom F, et al. Effectiveness of COVID-19 booster vaccines against COVID-19-related symptoms, hospitalization and death in England. Nat Med. 2022;28(4):831–837. doi:10.1038/s41591-022-01699-1

[10] Centers for Disease Control and Prevention. COVID Data Tracker. Accessed November 24, 2022. https://covid.cdc.gov/covid-data-tracker.

[11] Zhao S, Lou J, Cao L, et al. Differences in the case fatality risks associated with SARS-CoV-2 Delta and non-Delta variants in relation to vaccine coverage: An early ecological study in the United Kingdom. Infect Genet Evol. 2022;97(105162):105162. doi:10.1016/j.meegid.2021.105162

[12] Lee YC, Chang KY, Mirsaeidi M. Association of COVID-19 case-fatality rate with state health disparity in the United States. Front Med (Lausanne). 2022;9:853059. doi:10.3389/fmed.2022.853059

[13] Lipsitch M, Dean NE. Understanding COVID-19 vaccine efficacy. Science. 2020;370(6518):763–765. doi:10.1126/science.abe5938

[14] Halloran ME, Haber M, Longini IM Jr, Struchiner CJ. Direct and indirect effects in vaccine efficacy and effectiveness. Am J Epidemiol. 1991;133(4):323–331. doi:10.1093/oxfordjournals.aje.a115884

[15] Morens DM, Folkers GK, Fauci AS. The concept of classical herd immunity may not apply to COVID-19. J Infect Dis. 2022;226(2):195–198. doi:10.1093/infdis/jiac109

[16] Randolph HE, Barreiro LB. Herd immunity: Understanding COVID-19. Immunity. 2020;52(5):737–741. doi:10.1016/j.immuni.2020.04.012

[17] Gallagher ME, Sieben AJ, Nelson KN, et al. Indirect benefits are a crucial consideration when evaluating SARS-CoV-2 vaccine candidates. Nat Med. 2021;27(1):4–5. doi:10.1038/s41591-020-01172-x

[18] Flanagan BE, Hallisey EJ, Adams E, Lavery A. Measuring community vulnerability to natural and anthropogenic hazards: The centers for disease control and prevention’s social vulnerability index. J Environ Health. 2018;80(10):34–36.

[19] Dong E, Du H, Gardner L. An interactive web-based dashboard to track COVID-19 in real time. Lancet Infect Dis. 2020;20(5):533–534. doi:10.1016/S1473-3099(20)30120-1

[20] Place and Health, Agency for Toxic Substances and Disease Registry. CDC SVI 2018 Documentation; 2020. Accessed June 7, 2022. https://www.atsdr.cdc.gov/placeandhealth/svi/documentation/SVI_documentation_2018.html.

[21] Muggeo VMR. Estimating regression models with unknown breakpoints. Stat Med. 2003;22(19):3055–3071. doi:10.1002/sim.1545

[22] Muggeo VMR. Segmented: an R package to fit regression models with broken-line relationships. R news. 2008;8(1):20–25.

[23] McKee M, Rajan S. What can we learn from Israel’s rapid roll out of COVID 19 vaccination? Isr J Health Policy Res. 2021;10(5):1–4.

[24] Sah P, Vilches TN, Moghadas SM, et al. Accelerated vaccine rollout is imperative to mitigate highly transmissible COVID-19 variants. EClinicalMedicine. 2021;35(100865):100865. doi:10.1016/j.eclinm.2021.100865

[25] Moghadas SM, Sah P, Fitzpatrick MC, et al. COVID-19 deaths and hospitalizations averted by rapid vaccination rollout in the United States. bioRxiv. Published online 2021. doi:10.1101/2021.07.07.21260156

[26] Aguilera X, Mundt AP, Araos R, Weitzel T. The story behind Chile’s rapid rollout of COVID-19 vaccination. Travel Med Infect Dis. 2021;42(102092):102092. doi:10.1016/j.tmaid.2021.102092

[27] Glied S. Strategy drives implementation: COVID vaccination in Israel. Isr J Health Policy Res. 2021;10(1):9. doi:10.1186/s13584-021-00445-1

[28] Vilches TN, Moghadas SM, Sah P, et al. Estimating COVID-19 infections, hospitalizations, and deaths following the US vaccination campaigns during the pandemic. JAMA Netw Open. 2022;5(1):e2142725. doi:10.1001/jamanetworkopen.2021.42725

[29] Monge S, Olmedo C, Alejos B, et al. Direct and indirect effectiveness of mRNA vaccination against severe acute respiratory syndrome Coronavirus 2 in long-term care facilities, Spain. Emerg Infect Dis. 2021;27(10):2595–2603. doi:10.3201/eid2710.211184

[30] Clemente-Suárez VJ, Hormeño-Holgado A, Jiménez M, et al. Dynamics of population immunity due to the herd effect in the COVID-19 pandemic. Vaccines (Basel). 2020;8(2):236. doi:10.3390/vaccines8020236

[31] Brown CC, Young SG, Pro GC. COVID-19 vaccination rates vary by community vulnerability: A county-level analysis. Vaccine. 2021;39(31):4245–4249. doi:10.1016/j.vaccine.2021.06.038

[32] Chen Y, Ma ZF, Yu D, Jiang Z, Wang B, Yuan L. Geographical distribution of trace elements (selenium, zinc, iron, copper) and case fatality rate of COVID-19: a national analysis across conterminous USA. Environ Geochem Health. 2022;44(12):4423–4436. doi:10.1007/s10653-022-01204-0

[33] Weisel O. Vaccination as a social contract: The case of COVID-19 and US political partisanship. Proc Natl Acad Sci U S A. 2021;118(13):e2026745118. doi:10.1073/pnas.2026745118

[34] Li T, White LF. Bayesian back-calculation and nowcasting for line list data during the COVID-19 pandemic. PLoS Comput Biol. 2021;17(7):e1009210. doi:10.1371/journal.pcbi.1009210

[35] Bergen N, Kirkby K, Fuertes CV, et al. Global state of education-related inequality in COVID-19 vaccine coverage, structural barriers, vaccine hesitancy, and vaccine refusal: findings from the Global COVID-19 Trends and Impact Survey. Lancet Glob Health. 2023;11(2):e207–e217. doi:10.1016/S2214-109X(22)00520-4

[36] Chen Y, Zhang L, Li T, Li L. Amplified effect of social vulnerability on health inequality regarding COVID-19 mortality in the USA: the mediating role of vaccination allocation. BMC Public Health. 2022;22(1):2131. doi:10.1186/s12889-022-14592-w

[37] Buscot MJ, Wotherspoon SS, Magnussen CG, et al. Bayesian hierarchical piecewise regression models: a tool to detect trajectory divergence between groups in long-term observational studies. BMC Med Res Methodol. 2017;17(1). doi:10.1186/s12874-017-0358-9

[38] Li T. The Bayesian Paradigm of Robustness Indices of Causal Inferences. Michigan State University; 2018.

[39] Li T, Frank K. The probability of a robust inference for internal validity. Sociol Methods Res. 2022;51(4):1947–1968. doi:10.1177/0049124120914922

[40] Li T, Frank KA. The probability of a robust inference for internal validity and its applications in regression models. arXiv [statME]. Published online 2020. http://arxiv.org/abs/2005.12784

[41] Li T. On the probability of invalidating a causal inference due to limited external validity. arXiv [statME]. Published online 2022. http://arxiv.org/abs/2206.08649

