## Appendix for "Investigating the marginal and herd effects of COVID-19 vaccination for reducing case fatality rate: Evidence from the United States"

1-R code:

```
## Read the vaccination dataset
vacc <- read.csv('COVID-19_Vaccinations_in_the_United_States_County.csv')
## See how the dataset looks like for a single county
v1 <- vacc[vacc$FIPS==21151,]
## Read the raw time-series case counts
case <- read.csv('Raw Data/time_series_covid19_confirmed_US.csv')
## Read the raw time-series death counts
death <- read.csv('Raw Data/time_series_covid19_deaths_US.csv')
## Creat case fatality ratio
cfr <- death[,-(1:12)]/case[,-(1:11)]
cfr <- cbind.data.frame(death[,1:12],cfr)
write.csv(cfr,'cfr.csv',row.names = FALSE)

## Code for cleaning the data should be put here.
## The cleaned data is project.csv
## The data project.csv needs to be further cleaned, to address the following issues:
## Get rid of unknown counties.
## Remove rows with missing values in ppct, sppct and cfr.

## Find the breakpoints
library(segmented)
library(tidyverse)
project <- read.csv('project.csv')
project <- project[,-6]
## project <- project[!is.na(project$FIPS),]
colnames(project) <-
c("date","fips","county","state","ppct","theme1","theme2","theme3","theme4","cfr")
## But we need to select the right period,i.e., between March 11, 2021 and Jan 26, 2022
## The kickoff date of vaccination campaign in the US is March 11, 2021
## The date President Biden announce all Americans are eligible for vaccination
## The end date is Jan 26, 2022 when CDC announce that more than 99.9% of the virus
are Omicron
project <-
project%>%mutate(date=as.Date(date),cfr=100*cfr)%>%filter(date>=as.Date("2021-3-
11")&date<=as.Date("2022-1-26"))
mod <- lm(cfr~ppct+theme1+theme2+theme3+theme4,data = project)
project1 <-
project%>%group_by(date)%>%summarise(mcfr=mean(cfr),mppct=mean(ppct))
plot(project1$mppct,project1$mcfr,type='l')

## First analysis
## Segmented regression based on the mean data project1
```

```
## National level
mod <- lm(mcfr~mppct,data=project1)
selgmented(mod,seg.Z = ~mppct,type = "bic",Kmax = 3)
smod <- segmented(mod,seg.Z = ~mppct,npsi = 3)

## We identified 3 breakpoints,i.e.,32,36 and 47
plot(project1$mppct,project1$mcfr,type='l')
abline(v=32,lty=2,col='red')
abline(v=36,lty=2,col='red')
abline(v=47,lty=2,col='red')

## Create figure 1
k=data.frame(mppct=c(32,36,47),mcfr=c(1.97,1.758,1.767))
project1%>%ggplot(aes(x=mppct/100,y=mcfr/100))+geom_line()+geom_point(data=k)
+scale_y_continuous(
name = "Mean CFR", labels = scales::label_percent())+scale_x_continuous(
  name = "Mean Vaccination Coverage", labels =
scales::label_percent())+geom_point(data = k)

## We created the segmented regression model ourselves
project1 <-
project1%>%mutate(d1=ifelse(mppct>=32&mppct<36,1,0),d2=ifelse(mppct>=36&mp
pct<47,1,0),d3=ifelse(mppct>=47,1,0))
project1 <- project1%>%mutate(mppct1=d1*(mppct-32),mppct2=d2*(mppct-
36),mppct3=d3*(mppct-47))
mod1 <- lm(mcfr~mppct+d1+d2+d3+mppct1+mppct2+mppct3,data = project1)
summary(mod1)

## Calculate the p-values associated with the herd and marginal effects
m=vcov(mod1)
coe=coef(mod1)
## herd2
v=c(0,0,-1,1,0,0,0,0)
2*pt(sum(coe*v)/sqrt(t(v)%*%m%*%v),314)
## herd3
v=c(0,0,0,-1,1,0,0,0)
2*(1-pt(sum(coe*v)/sqrt(t(v)%*%m%*%v),314))
## margin2
v=c(0,1,0,0,0,1,0,0)
2*pt(sum(coe*v)/sqrt(t(v)%*%m%*%v),314)
## margin3
v=c(0,1,0,0,0,0,1,0)
2*(1-pt(sum(coe*v)/sqrt(t(v)%*%m%*%v),314))
## margin4
```

```
v=c(0,1,0,0,0,0,0,1)
2*pt(sum(coe*v)/sqrt(t(v)%*%m%*%v),314)

## Run mixed models
## First create dummy variables based on the three breakpoints
project <- project%>%mutate(d1=ifelse(ppct>=32&ppct<36,1,0),d2=ifelse(ppct>=36&ppct<47,1,0),d3=ifelse(ppct>=47,1,0))
project <- project%>%mutate(ppct1=d1*(ppct-32),ppct2=d2*(ppct-36),ppct3=d3*(ppct-47))
## See an example
dat <- project%>%filter(fips==6113)
mod <- lm(cfr~ppct+d1+d2+d3+ppct1+ppct2+ppct3,data = dat)
summary(mod)
plot(dat$ppct,dat$cfr,type='l')

## Use lmer function
library(lme4)
library(RLRsim)
mixmod0 <- lmer(cfr~1+ppct+d1+d2+d3+ppct1+ppct2+ppct3+(1|fips)+theme1+theme2+theme3+theme4,data = project,REML=FALSE,control=lmerControl(optimizer="bobyqa",calc.derivs = FALSE,optCtrl=list(maxfun=2e5)))
mixmod1 <- lmer(cfr~1+ppct+d1+d2+d3+ppct1+ppct2+ppct3+(1+ppct+ppct1+ppct2+ppct3|fips)+theme1+theme2+theme3+theme4,data = project,REML=FALSE,control=lmerControl(optimizer="bobyqa",calc.derivs = FALSE,optCtrl=list(maxfun=2e5)))
mixmod2 <- lmer(cfr~1+ppct+d1+d2+d3+ppct1+ppct2+ppct3+(1+ppct+d1+d2+d3+ppct1+ppct2+ppct3|fips)+theme1+theme2+theme3+theme4,data = project,REML=FALSE,control=lmerControl(optimizer="bobyqa",calc.derivs = FALSE,optCtrl=list(maxfun=2e5)))

## Running the code above (about mixed models) was quite slow in R.
## So we used STATA to run the same models, see details in the appendix.

## Calculate the p-values associated with the herd and marginal effects
## herd2
tst=(-0.0175721+0.0251904)/(0.00001174+0.0003834-2*6.209e-07)^0.5
2*(1-pt(tst,310))
## herd3
tst=(-0.0039231+0.0175721)/(0.0001276+0.0003834-2*2.033e-06)^0.5
```

```
2*(1-pt(tst,310))
## margin2
tst=(-0.0036004-0.0067726)/(1.654e-07+3.284e-06-2*4.495e-09)^0.5
2*pt(tst,310)
## margin3
tst=(-0.0036004-0.0196573)/(1.654e-07+2.261e-06-2*4.983e-09)^0.5
2*pt(tst,310)
## margin4
tst=(-0.0036004-0.0393751)/(1.654e-07+2.809e-06-2*5.985e-09)^0.5
2*pt(tst,310)
```

### 2-Analysis at county level (mixed model) using STATA:

#### 1. model1: random effects for all terms except theme1 through theme4

```
. mixed c.cfr c.vc i.d1 i.d2 i.d3 c.vc1 c.vc2 c.vc3 theme1 theme2 theme3 theme4 || fips: c.vc i.d1 i.d2 i.d3 c
> .vc1 c.vc2 c.vc3, mle
```

**Output:**

```
Mixed-effects ML regression                     Number of obs     =  1,001,098
Group variable: fips                            Number of groups  =      3,109
                                                Obs per group:
                                                              min =        322
                                                              avg =      322.0
                                                              max =        322
                                                Wald chi2(11)     =    1016.31
Log likelihood =  513590.06                     Prob > chi2       =     0.0000
```

| cfr | Coefficient | Std. err. | z | P>\|z\| | [95% conf. | interval] |
|---|---|---|---|---|---|---|
| vc | -.0036004 | .0004067 | -8.85 | 0.000 | -.0043975 | -.0028034 |
| 1.d1 | -.0251904 | .0034266 | -7.35 | 0.000 | -.0319065 | -.0184743 |
| 1.d2 | -.0175721 | .0061915 | -2.84 | 0.005 | -.0297073 | -.0054369 |
| 1.d3 | -.0039231 | .0112962 | -0.35 | 0.728 | -.0260633 | .0182171 |
| vc1 | -.0067726 | .0018122 | -3.74 | 0.000 | -.0103244 | -.0032208 |
| vc2 | -.0196573 | .0015037 | -13.07 | 0.000 | -.0226045 | -.01671 |
| vc3 | -.0393751 | .0016761 | -23.49 | 0.000 | -.0426603 | -.0360899 |
| theme1 | .0003766 | .0011555 | 0.33 | 0.744 | -.0018882 | .0026414 |
| theme2 | .8068916 | .0733619 | 11.00 | 0.000 | .6631049 | .9506783 |
| theme3 | -.0573455 | .0788306 | -0.73 | 0.467 | -.2118507 | .0971597 |
| theme4 | .0038997 | .0806247 | 0.05 | 0.961 | -.1541218 | .1619213 |
| _cons | 1.640176 | .0567852 | 28.88 | 0.000 | 1.528879 | 1.751473 |

| Random-effects parameters | Estimate | Std. err. | [95% conf. | interval] |
|---|---|---|---|---|
| fips: Independent | | | | |
| var(vc) | .0004989 | .0000129 | .0004743 | .0005248 |
| var(1.d1) | .0238676 | .0007884 | .0223713 | .0254639 |
| var(1.d2) | .0912826 | .0028374 | .0858875 | .0970167 |
| var(1.d3) | .1991367 | .0074995 | .1849673 | .2143915 |
| var(vc1) | .0072339 | .000239 | .0067802 | .0077178 |
| var(vc2) | .0056307 | .0001856 | .0052784 | .0060064 |
| var(vc3) | .0042457 | .0001783 | .0039102 | .0046099 |
| var(_cons) | 1.32611 | .0338076 | 1.261477 | 1.394055 |
| var(Residual) | .0189058 | .000027 | .0188529 | .0189588 |

```
LR test vs. linear model: chi2(8) = 3.6e+06                Prob > chi2 = 0.0000

Note: LR test is conservative and provided only for reference.
```

The covariance of random effects

```
. estat recovariance
```

Random-effects covariance matrix for level **fips**

| | d1 | d2 | d3 | ppct | ppct1 | ppct2 |
|---|---|---|---|---|---|---|
| d1 | .0238676 | | | | | |
| d2 | 0 | .0912826 | | | | |
| d3 | 0 | 0 | .1991367 | | | |
| ppct | 0 | 0 | 0 | .0004989 | | |
| ppct1 | 0 | 0 | 0 | 0 | .0072339 | |
| ppct2 | 0 | 0 | 0 | 0 | 0 | .0056307 |
| ppct3 | 0 | 0 | 0 | 0 | 0 | 0 |
| _cons | 0 | 0 | 0 | 0 | 0 | 0 |

| | ppct3 | _cons |
|---|---|---|
| ppct3 | .0042457 | |
| _cons | 0 | 1.32611 |

The covariance matrix of the fixed effects in model 1:

```
. estat vce
```

Covariance matrix of coefficients of **mixed** model

| e(V) | cfr<br>vc | 1.<br>d1 | 1.<br>d2 | 1.<br>d3 | vc1 | vc2 | vc3 |
|---|---|---|---|---|---|---|---|
| **cfr** | | | | | | | |
| vc | 1.654e-07 | | | | | | |
| 1.d1 | -2.852e-08 | .00001174 | | | | | |
| 1.d2 | -7.419e-08 | 6.209e-07 | .00003834 | | | | |
| 1.d3 | -1.566e-07 | 1.011e-06 | 2.033e-06 | .0001276 | | | |
| vc1 | -4.495e-09 | -7.162e-07 | 9.894e-08 | 1.485e-07 | 3.284e-06 | | |
| vc2 | -4.983e-09 | 2.775e-08 | -2.885e-07 | 1.843e-07 | 4.746e-09 | 2.261e-06 | |
| vc3 | -5.985e-09 | 3.483e-08 | 6.949e-08 | -1.201e-07 | 5.876e-09 | 6.619e-09 | 2.809e-06 |

Save this model as m1 in STATA.

```
. est sto m1
```

2. We noticed that the random effect sizes of vc, vc1, vc2, vc3 were small, and therefore we considered removing the random effects of those four terms in the model2.

```
. mixed c.cfr c.vc i.d1 i.d2 i.d3 c.vc1 c.vc2 c.vc3 theme1 theme2 theme3 theme4
> || fips: i.d1 i.d2 i.d3, mle
```

Output:

```
Iteration 0:   log likelihood =   209406.2
Iteration 1:   log likelihood =   209406.2

Computing standard errors ...

Mixed-effects ML regression                     Number of obs     =  1,001,098
Group variable: fips                            Number of groups  =      3,109
                                                Obs per group:
                                                              min =        322
                                                              avg =      322.0
                                                              max =        322
                                                Wald chi2(11)     =   84270.90
Log likelihood =   209406.2                     Prob > chi2       =     0.0000

------------------------------------------------------------------------------
         cfr | Coefficient  Std. err.      z    P>|z|     [95% conf. interval]
-------------+----------------------------------------------------------------
          vc |  -.0058018   .0000486  -119.40   0.000     -.005897   -.0057065
        1.d1 |   .0042839   .0056506     0.76   0.448    -.0067912     .015359
        1.d2 |   .0395135   .0067861     5.82   0.000     .0262129     .052814
        1.d3 |   .0471648   .0103839     4.54   0.000     .0268126    .0675169
         vc1 |  -.0037126   .0005969    -6.22   0.000    -.0048825   -.0025427
         vc2 |  -.0112739   .0001549   -72.77   0.000    -.0115776   -.0109703
         vc3 |  -.0150956   .0001016  -148.56   0.000    -.0152948   -.0148965
      theme1 |   .0002682   .0009363     0.29   0.775     -.001567    .0021034
      theme2 |    .882877    .059442    14.85   0.000     .7663728    .9993812
      theme3 |  -.0280334   .0639139    -0.44   0.661    -.1533022    .0972355
      theme4 |  -.0121168   .0653133    -0.19   0.853    -.1401285    .1158948
       _cons |   1.643514   .0460278    35.71   0.000     1.553301    1.733727
------------------------------------------------------------------------------

------------------------------------------------------------------------------
  Random-effects parameters  |   Estimate   Std. err.     [95% conf. interval]
-----------------------------+------------------------------------------------
fips: Independent            |
                   var(1.d1) |   .0751658   .0021548      .0710589    .0795101
                   var(1.d2) |   .1188571   .0032836      .1125925    .1254703
                   var(1.d3) |   .1795017   .0062097      .1677343    .1920947
                  var(_cons) |   .8738916   .0221827      .8314781    .9184686
-----------------------------+------------------------------------------------
               var(Residual) |   .0362979   .0000516       .036197    .0363992
------------------------------------------------------------------------------
LR test vs. linear model: chi2(4) = 3.0e+06           Prob > chi2 = 0.0000

Note: LR test is conservative and provided only for reference.
```

Save this model as m2 in STATA.

```
. est sto m2
```

#### 3. Using Likelihood Ratio Test to compare m1 and m2.

```
. lrtest m1 m2, stats

Likelihood-ratio test
Assumption: m2 nested within m1

 LR chi2(4) = 608367.73
Prob > chi2 =    0.0000

Note: The reported degrees of freedom assumes the null hypothesis is not on
      the boundary of the parameter space. If this is not true, then the
      reported test is conservative.

Akaike's information criterion and Bayesian information criterion
```

| Model | N | ll(null) | ll(model) | df | AIC | BIC |
|---|---|---|---|---|---|---|
| m2 | 1,001,098 | . | 209406.2 | 17 | -418778.4 | -418577.5 |
| m1 | 1,001,098 | . | 513590.1 | 21 | -1027138 | -1026890 |

Note: BIC uses N = number of observations. See [R] BIC note.

We rejected the null hypothesis and concluded that m1 was better than m2.

4. **Furthermore, we observed that the random effect size of vc was the smallest. Therefore, we only removed the random effect of vc in the model3 and save this model as m3 in STATA.**

```
Performing EM optimization ...

Performing gradient-based optimization:
Iteration 0:   log likelihood =  322562.17
Iteration 1:   log likelihood =  322562.17

Computing standard errors ...

Mixed-effects ML regression                     Number of obs     =  1,001,098
Group variable: fips                            Number of groups  =      3,109
                                                Obs per group:
                                                              min =        322
                                                              avg =      322.0
                                                              max =        322
                                                Wald chi2(11)     =   19757.01
Log likelihood =  322562.17                     Prob > chi2       =     0.0000

------------------------------------------------------------------------------
         cfr | Coefficient  Std. err.      z    P>|z|     [95% conf. interval]
-------------+----------------------------------------------------------------
          vc |  -.0057919   .0000429  -135.01   0.000     -.005876   -.0057078
        1.d1 |   .0003946   .0047291     0.08   0.934    -.0088743    .0096634
        1.d2 |   .0289429   .0066543     4.35   0.000     .0159007    .0419852
        1.d3 |   .0863778   .0097953     8.82   0.000     .0671793    .1055763
         vc1 |  -.0045298   .0018739    -2.42   0.016    -.0082025   -.0008571
         vc2 |  -.0151826   .0013547   -11.21   0.000    -.0178378   -.0125274
         vc3 |  -.0336915   .0015201   -22.16   0.000    -.0366709    -.030712
      theme1 |   .0002685   .0009378     0.29   0.775    -.0015696    .0021066
      theme2 |   .8856445   .0595413    14.87   0.000     .7689456    1.002343
      theme3 |  -.0279839   .0640191    -0.44   0.662     -.153459    .0974912
      theme4 |  -.0143341   .0654228    -0.22   0.827    -.1425604    .1138923
       _cons |   1.642384   .0461006    35.63   0.000     1.552028     1.73274
------------------------------------------------------------------------------

------------------------------------------------------------------------------
  Random-effects parameters  |   Estimate   Std. err.     [95% conf. interval]
-----------------------------+------------------------------------------------
fips: Independent            |
                  var(1.d1)  |   .0490266   .0014968      .0461791    .0520498
                  var(1.d2)  |   .1100243   .0032219      .1038872     .116524
                  var(1.d3)  |   .1573158   .0056196      .1466784    .1687247
                   var(vc1)  |   .0073673    .000241      .0069097    .0078551
                   var(vc2)  |   .0044818   .0001504      .0041965    .0047865
                   var(vc3)  |   .0034149   .0001469      .0031388    .0037152
                 var(_cons)  |   .8768629   .0222595      .8343026    .9215942
-----------------------------+------------------------------------------------
              var(Residual)  |   .0282662   .0000403      .0281873    .0283453
------------------------------------------------------------------------------
LR test vs. linear model: chi2(7) = 3.2e+06           Prob > chi2 = 0.0000

Note: LR test is conservative and provided only for reference.

.  est sto m3
```

### 5. Use Likelihood Ratio Test to compare m1 and m3.

```
. lrtest m1 m3, stats

Likelihood-ratio test
Assumption: m3 nested within m1

 LR chi2(1) = 382055.78
Prob > chi2 =    0.0000

Note: The reported degrees of freedom assumes the null hypothesis is not on the boundary of the parameter
      space. If this is not true, then the reported test is conservative.

Akaike's information criterion and Bayesian information criterion
```

| Model | N | ll(null) | ll(model) | df | AIC | BIC |
|---|---|---|---|---|---|---|
| m3 | 1,001,098 | . | 322562.2 | 20 | -645084.3 | -644848 |
| m1 | 1,001,098 | . | 513590.1 | 21 | -1027138 | -1026890 |

```
Note: BIC uses N = number of observations. See [R] BIC note.
```

We rejected the null hypothesis and concluded that m1 was still better than m3.
